# Unlocking the Code: A Qualitative Study on what Drives or Deters South African Adolescents to Embrace Clinical Research?

**DOI:** 10.1101/2025.04.13.25325756

**Authors:** Yvonne Wangui Machira, Heeran Makkan, Funeka Mthembu, Pholo Maenetje, Matt A. Price, Vinodh Edward, Candice Chetty-Makkan

**Affiliations:** IAVI Africa, Nairobi, Kenya; The Aurum Institute, Rustenburg Clinical Research Centre, Rustenburg, South Africa; Health Economics and Epidemiology Research Office (HE2RO), Faculty of Health Sciences, Wits Health Consortium, University of the Witwatersrand, South Africa; IAVI, New York City, New York, United States of America; Department of Epidemiology and Biostatistics, University of California at San Francisco, San Francisco, California, United States of America

## Abstract

**Background:** Understanding adolescent decision-making processes is crucial for designing effective clinical trials that cater to their unique needs and perspectives. Adolescents are at high risk for contracting HIV where their behaviours, perceptions and attitudes could influence treatment responses and long-term health outcomes. Therefore, integrating adolescent perspectives is critical in clinical research, yet, adolescent perspectives remain underrepresented in clinical trials. This study addresses this gap by examining the drivers and barriers to clinical research participation among adolescents and their parents/guardians in Rustenburg, South Africa.

**Methods:** We conducted 6 focus group discussions (FGDs) with 55 participants consisting of separate sessions for 36 adolescents (12-19 years) and 19 parents/guardians to ensure open dialogue within each group. At baseline (April to July 2018), we conducted two FGDs with adolescents (N=17) and two FGDs with parents/guardians (N=19). At endline (August to November 2019), we conducted an additional two FGDs with a different group of adolescents (N=19). Conducting FGDs at these two timepoints helped identify attributes before and after trial participation. Thematic analysis guided by the COM-B (Capability, Opportunity, Motivation - Behaviour) framework was used to examine factors influencing adolescent participation in clinical research in Rustenburg. The analysis used both deductive coding based on COM-B components and inductive approaches to capture additional themes. Using QSR NVIVO 12 and 14 qualitative analysis software, we conducted iterative coding of FGD transcripts and debrief notes to identify, categorise, and refine themes related to capabilities (knowledge and skills), opportunities (physical and social environment), and motivations (reflective and automatic) that influence research participation decisions.

**Results:** We conducted FGDs with 36 adolescents (24 females, 12 males; median age 17.5 years, IQR 15-18) and 19 parents/guardians (17 females, 2 males; median age 37 years, IQR 34-46). Analysis using the COM-B framework revealed how capability, opportunity, and motivation influenced research participation. Psychological capability improved through knowledge acquisition, with adolescents demonstrating enhanced understanding of clinical research from baseline to endline. Physical and social opportunities manifested through access to healthcare services and community engagement, with community leaders’ endorsement playing a pivotal role. Reflective motivation emerged through altruism, while automatic motivation was evident in emotional responses. Barriers included limited psychological capability (lack of awareness about research processes), restricted physical opportunities (logistical challenges with transportation and school schedules), and negative automatic motivations (fear and mistrust). Initial high levels of fear decreased by endline, while mistrust rooted in past experiences and community narratives remained persistent. These themes emerged consistently across baseline and endline FGDs, with variations between adolescents and parents/guardians.

**Conclusions:** Our results highlight the importance of integrating adolescent preferences when designing clinical trials to enhance enrolment and retention. Analysed through the COM-B framework, the study’s cross-sectional design revealed that participation itself can increase capability (knowledge and awareness) and motivation (trust) over time. Researchers should prioritize effective, age-appropriate communication strategies, community engagement, and flexible logistics to address identified barriers to capability, opportunity, and motivation. These insights can guide the development of more inclusive and supportive research environments, particularly crucial in South Africa with its large youth population.

## Introduction

In recent years, the importance of involving adolescents in clinical research has gained significant attention (1). Adolescents, a population characterised by rapid physical, emotional, and cognitive changes, present unique health challenges and opportunities (2). Understanding adolescent health dynamics is pivotal (3), especially in regions like Rustenburg, South Africa, where individual, social, and contextual play a crucial role in health outcomes.

The participation of adolescents in clinical research is not just a matter of inclusivity - it is a necessity (4). This is because adolescents’ physiological and psychological differences from adults can lead to different disease manifestations, treatment responses, and long-term health outcomes (2). Despite this, globally, the adolescent demographic remains underrepresented in clinical trials, leading to a knowledge gap that hinders the development of targeted interventions (5).

Globally, efforts to increase adolescent participation in clinical trials have been gaining momentum. Over the past two decades, there has been a growing recognition of the importance of including adolescents in medical research, particularly in areas such as HIV prevention, mental health, and reproductive health. Many countries have updated their regulatory frameworks to facilitate adolescent participation while ensuring appropriate protections are in place. For example, the European Medicines Agency (EMA) released guidelines in 2017 specifically addressing the inclusion of adolescents, classified as paediatrics, in clinical trials (6). In the United States, the National Institutes of Health (NIH) mandated the inclusion of children in research in 1998 (7), leading to increased adolescent participation across various fields (4). However, challenges remain. A 2023 study (8) found that many regulatory guidance documents are silent on or exclusionary towards adolescent inclusion in adult trials. Their analysis of Food and Drug Administration (FDA) and EMA guidance documents revealed that only 32 per cent of FDA and 15 per cent of EMA documents contained permissive language for adolescent inclusion (8). The study highlighted the need for more explicit recommendations on age-inclusive trial designs in regulatory guidance (8). Additionally, regulatory bodies in African countries have made significant strides in developing frameworks to support ethical adolescent research participation; for instance, South Africa has implemented comprehensive guidelines that balance protecting adolescent welfare with recognising their capacity for informed decision-making (12).

In Africa, these efforts have been particularly focused on HIV research, given the high burden of the disease among young people in the region. The UNAIDS 2018 report highlighted that adolescents (15-24 years) accounted for 36 per cent of new HIV infections in Eastern and Southern Africa (ESA) (9)(10). Over the last six years, this has led to increased efforts to include adolescents in HIV prevention trials, including vaccine studies and pre-exposure prophylaxis (PrEP) research. Countries like South Africa, Kenya, and Uganda have been at the forefront of these efforts, developing specific ethical guidelines for adolescent participation in HIV research (11). Additionally, there’s been a growing emphasis on addressing other health issues affecting African youth, such as malaria, tuberculosis, and sexual and reproductive health, through adolescent-inclusive research approaches (3).

Despite these global and regional trends, significant challenges remain in terms of ethical considerations, consent processes, and designing adolescent-friendly research protocols, particularly in resource-limited settings common in many African countries. In South Africa, where societal norms and cultural beliefs play a significant role in health-seeking behaviours, and where youth-friendly services have emerged as a critical approach to adolescent healthcare, understanding the motivators and barriers to clinical research participation becomes even more crucial (12).

Existing literature has highlighted various drivers of adolescent participation in clinical research. These include knowledge acquisition, access to services, and community engagement (13). The role of parents/guardians in facilitating adolescent participation is also crucial, as their understanding and support can significantly influence adolescents’ willingness to engage in research (14,15).

However, several barriers to adolescents’ participation in clinical research have also been identified. These include fear, mistrust, logistical issues, and lack of awareness or understanding of the research process (13). Further, collecting accurate data on sensitive topics such as sexual behaviour can be problematic as we observed in the clinic where we conducted the work reported here (16). Additionally, parental/guardian concerns and misconceptions can further deter adolescent involvement (13)(14)(17,18). These barriers are often intertwined with ethical considerations surrounding adolescent participation in research (4)(19). While some argue that adolescents are incapable of providing informed consent due to their cognitive immaturity (20), others believe that with proper safeguards, their inclusion is both ethical and necessary (4).

This study was designed with the primary aim of delving into these dynamics, focusing on both the adolescents and their parents/guardians within the South African context. Through this research, we sought to provide insights that could guide the design of adolescent-centric clinical trials and health interventions in similar settings.

## Methods

### Research Design Overview

This study was nested within the Youth Friendly Services (YFS) 12-month cohort study that was conducted with 223 adolescents in Rustenburg from April 2018 to November 2020 (16). We employed a qualitative research design, focusing on data-collection strategies that included focus group discussions (FGDs) and field debrief notes. The data-analytic strategies encompassed thematic analysis, grounded theory, and narrative analysis. Our approach to inquiry was primarily interpretive, aiming to understand the underlying motivations, perceptions, and experiences of the participants. The rationale behind this design was to capture the richness and depth of participants’ perspectives, allowing for a comprehensive understanding of the drivers and barriers to clinical research participation among adolescents in Rustenburg.

### Research Team

The research team comprised individuals with diverse backgrounds in clinical research, adolescent health, and qualitative methodologies. To ensure objectivity and reduce potential biases, the team engaged in regular debriefing sessions, discussing interpretations and reflections throughout the research process. This approach helped manage researchers’ prior understandings, ensuring they enhanced rather than limited the data collection and analysis.

### Participant Selection

We selected participants to ensure maximum variation and capture a wide range of experiences and perspectives. For each participant group, parents/guardians of minor participants were required to provide written consent for their adolescent’s participation in the focus group discussions (FGDs). The inclusion criteria for adolescents included being 12 to 19 years old, residents of Rustenburg, willing and able to provide written assent (if under 18 years) or written consent (if 18 years and above), willing to participate in HIV testing and counselling, answer HIV and sexual behaviour questions, undergo a full physical examination, have a negative HIV status, and for females, have a negative pregnancy test. Though we conducted some interviews at baseline, and some at the end of follow up (after 9-12 months of follow up), no participant was interviewed twice. We planned to conduct at least two FGDs with parents/guardians and four with adolescents.

### Data Collection

The primary form of data collection was FGDs, with demographic data collected in the main YFS study. Using an FGD guide, and guided by a moderator, these discussions were designed to gather in-depth insights from both adolescents and their parents or guardians regarding their perceptions, experiences, and attitudes towards clinical research participation. We purposefully conducted 4 FGDs at baseline (2 with adolescents and 2 with parents/guardians), to describe attitudes at the project’s inception. A further 2 FGDs with adolescents at endline with a different group of participants, a year after the baseline discussions to describe how these attitudes may have evolved after study follow up.. On average, FGDs were held for approximately two hours, with a range of approximately one and half hours to two and a half hours, including a short break. This allowed ample time for participants to share their views, experiences, and suggestions while also ensuring that the discussions remained focused and relevant. Each FGD had an average of 9 participants.

The FGDs were conducted in the language preference of the participants, both English and Setswana, audio recordings and process notes were transcribed and where necessary translated prior to analysis. To ensure completeness of the notes and recordings, debriefing sessions were conducted between moderator and scribe to compare findings and address any new topics that arose during the FGDs.

The process of translations and transcriptions started within seven days of conduct of the FGDs. The FGDs that were conducted in English were transcribed verbatim. For FGDs that were conducted in Setswana, they were first transcribed verbatim and then transcribed into English. For quality control purposes, random checks between the audio recordings and transcriptions were conducted by an independent staff member.

The questions posed during the FGDs were open-ended, encouraging participants to share their thoughts freely. Some of the central questions included: (1) “*What are your perceptions about participating in clinical research?*”, (2) “*Have you or your child ever participated in a clinical trial? If so, what was the experience like?*”, (3) “*What do you believe are the main barriers to adolescents’ participation in clinical research?*”, (4) “*How do you think clinical trials can be made more adolescent-friendly?*” and (5) “*What motivates you or would motivate you to participate in clinical research?*”.

### Data Analysis

We employed thematic analysis as our primary analytical approach, allowing us to explore deeply into both the focus group transcripts and debriefing notes to uncover themes that best represented participants’ experiences, perceptions, and attitudes towards clinical research participation. Our analysis was conducted using QSR NVIVO 12 and 14 qualitative analysis software, following a systematic process that combined both deductive and inductive approaches to theme development.

To structure our analysis, we employed a systematic and iterative approach to codebook development. This process integrated focus group discussion (FGD) transcripts and debriefing notes as primary data sources. Initially, the research team conducted a comprehensive review of all data to identify potential themes, informed both by the research questions and emergent patterns within the dataset. Preliminary codes were developed and refined iteratively, with constant comparison and alignment to the theoretical framework guiding our study. These codes were organized hierarchically into main themes and sub-themes, ensuring coherence and relevance across the data.

To enhance analytical rigor and validity, multiple researchers (CCM, HM, and YWM) independently coded each transcript and corresponding debriefing notes. Discrepancies in coding were resolved through structured team discussions, fostering consensus and ensuring consistency. This collaborative approach also served to mitigate individual researcher biases. Themes were reviewed and validated against the entire dataset, with debriefing notes used to contextualize and corroborate interpretations of FGD discussions.

Throughout the data collection and analysis, we practiced reflexivity by continually reflecting on our positionality and potential biases. This included documenting personal assumptions and engaging in critical discussions within the team to ensure that the voices of participants remained central to the interpretation of findings (21,22)(23).

Our analysis was guided by the COM-B framework (24) which provides a theoretical structure for understanding behaviour change through three interacting components:

- Capability (psychological and physical): Knowledge and skills needed to participate in research
- Opportunity (social and physical): Factors in the environment that enable or prevent participation
- Motivation (reflective and automatic): Automatic and reflective mechanisms that activate or inhibit participation

The emergent themes from our thematic analysis were organised within this framework to understand how these components influence adolescents’ decisions about research participation.

### Thematic Framework

The thematic framework was primarily informed by a comprehensive analysis of multiple data sources. These sources encompassed a range of qualitative data, including debriefing notes, and transcripts from FGDs.

The analysis of these data sources was conducted using a systematic approach, ensuring that all relevant data points were captured. The data was then categorised and organised into themes and sub-themes, leading to the development of the thematic framework. The framework was designed to capture the complexities and nuances of adolescents’ participation in clinical research, especially in the specific context of Rustenburg, South Africa. The themes and sub-themes were derived directly from the data, ensuring that the framework was grounded in the actual experiences and perspectives of the participants.

The thematic framework revolves around the central theme of “*Adolescents’ Participation in Clinical Research*”. This framework identifies two main sub-themes: “*Motivators*” and “*Barriers*”. Each sub-theme further categorises the factors influencing adolescent participation in clinical research. The detailed explanation of these themes and sub-themes is presented in Table 1.

**Table 1:**
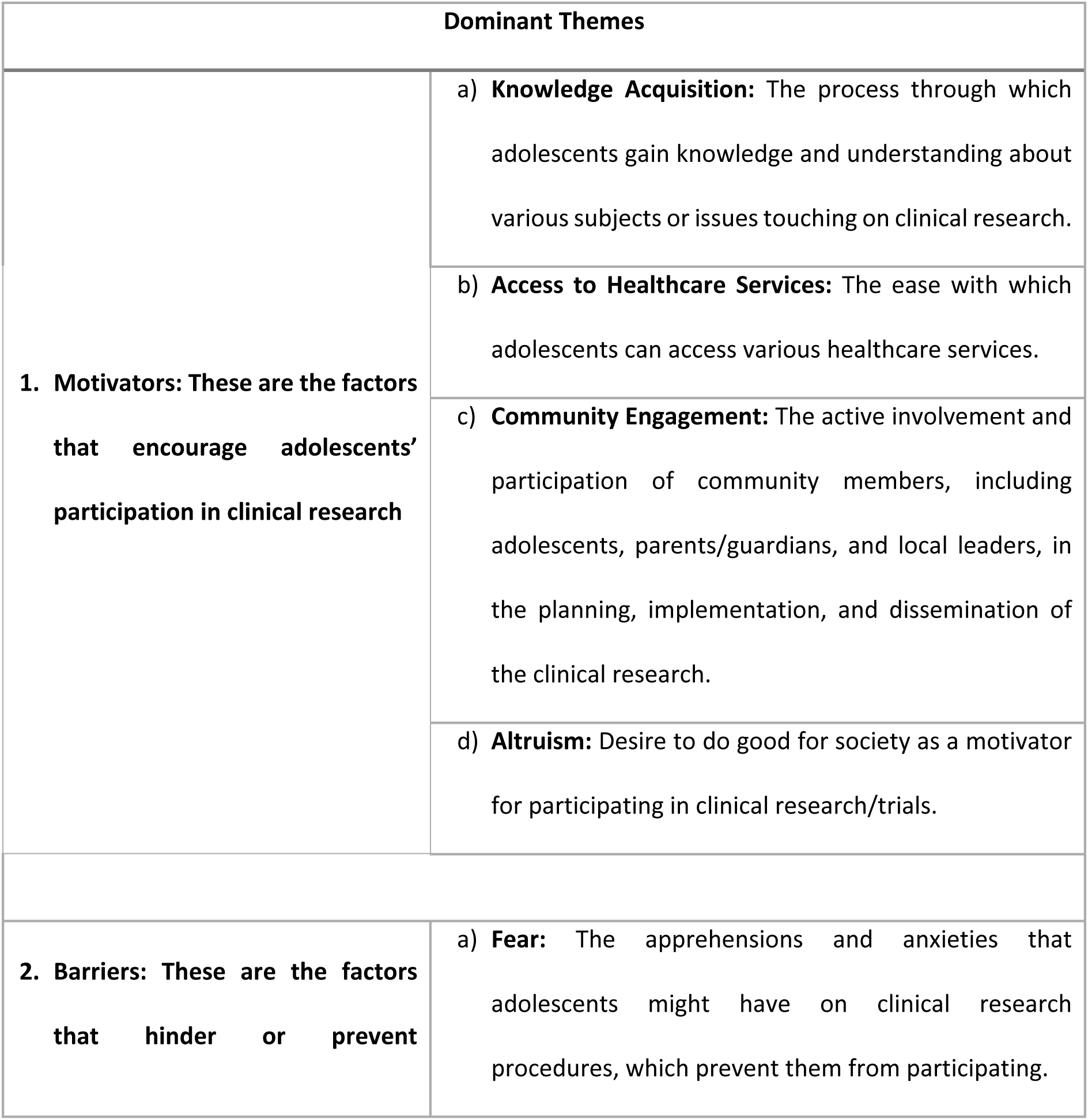
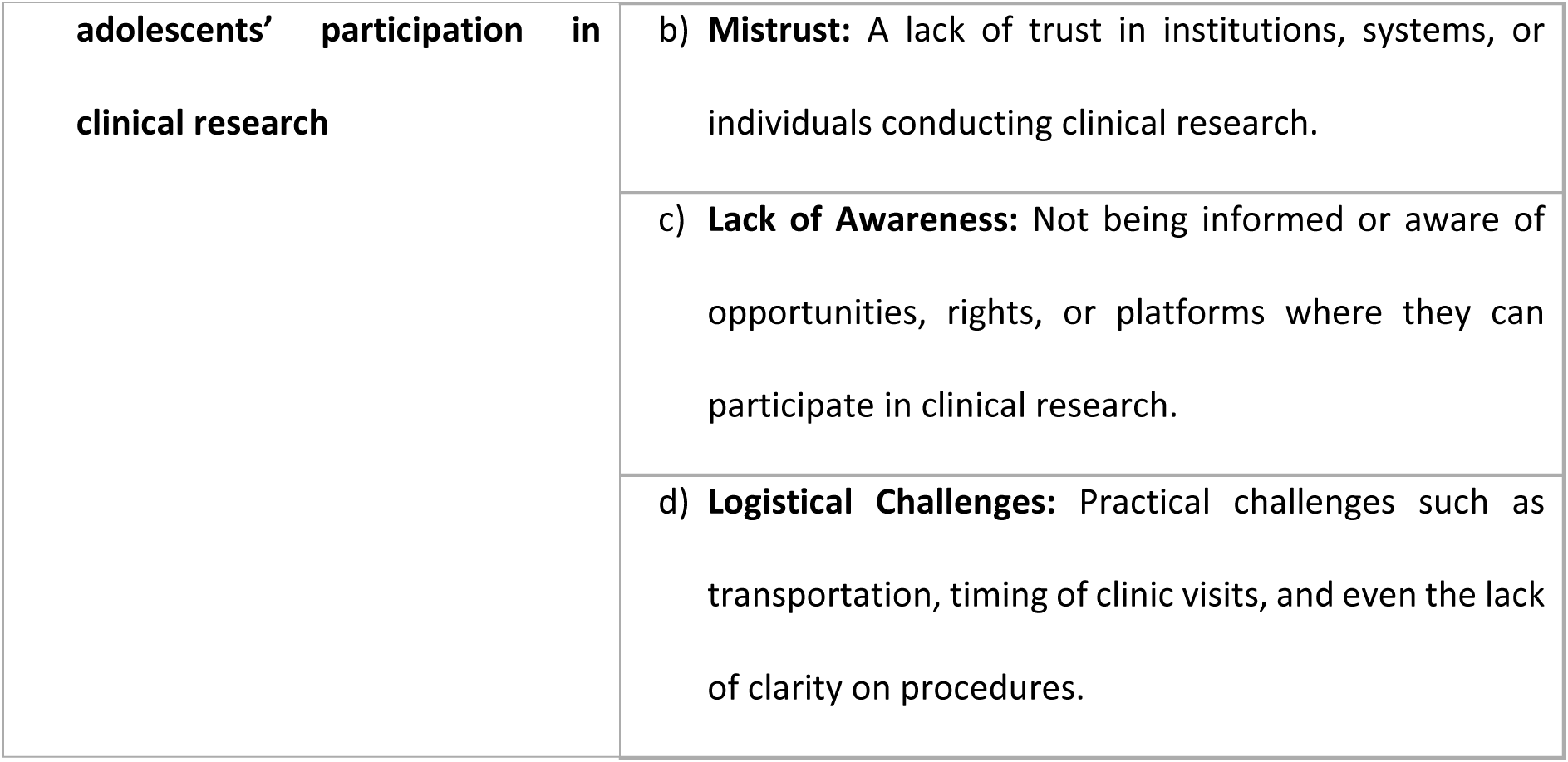
Explanation of Themes & Sub-Themes.

The emergent themes were then organised using the COM-B framework (24), which provides a theoretical structure for understanding behaviour change through three interacting components. Capability was primarily psychological, encompassing knowledge acquisition and lack of awareness and understanding about the research processes. Opportunity manifested in both physical forms (access to healthcare services and logistical challenges) and social dimensions (community engagement and support from research teams). Motivation included both reflective processes (altruism) and automatic responses (fear and mistrust).

This established framework was selected as it effectively captures the multiple factors influencing adolescents’ decisions to participate in clinical research, while allowing us to preserve the contextual richness of our findings. The framework enabled us to reconceptualize our original thematic framework - which separated motivators (knowledge acquisition, access to healthcare services, community engagement, and altruism) and barriers (fear, mistrust, lack of awareness, and logistical challenges) - into an integrated behavioural model that better explains how these factors interact to influence participation decisions as shown in the transition from the original framework to COM-B components in Figure 1.

**Figure 1:**
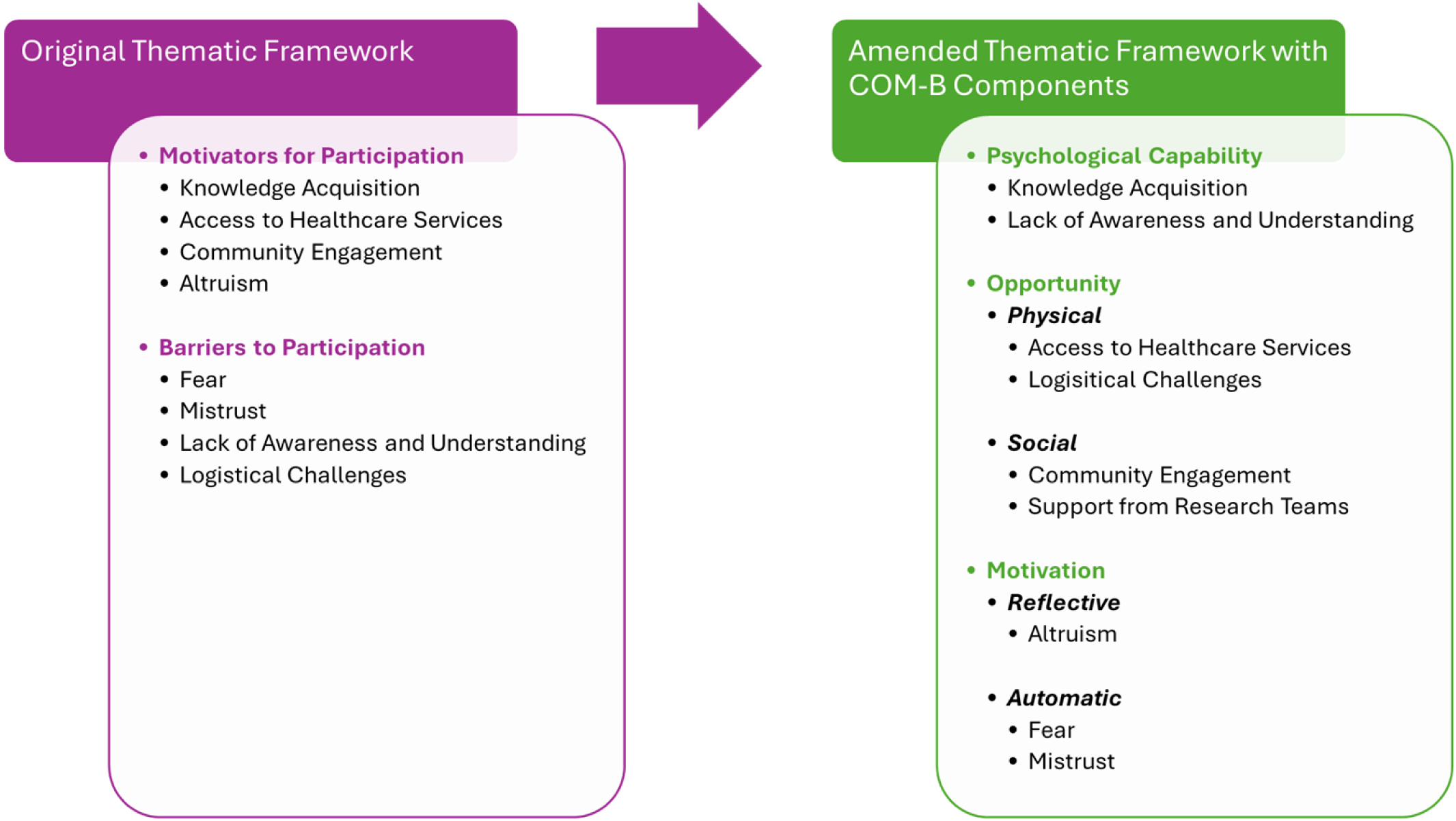
Thematic Framework of Adolescent Clinical Research Participation Using the COM-B Framework (24).

### Ethical Statement

The protocol, informed consent forms, assessment of understanding, other requested documents, and any subsequent modifications were reviewed and approved by, the University of Witwatersrand Human Research Ethics committee (Ref no. 170607). The Aurum Institute was also registered with the Office of Human Research Protection with the assurance number: FWA00004781.

Parent/legal guardians of minor participants (12-17 years old) who enrolled in the study were invited to provide parent/legal guardian written consent for their adolescent minors participated in the FGDs. The adolescent minors were also invited to provide written assent. Adolescents who were between 18 and 19 years old were invited to provide written consent. Participants were reimbursed ZAR150.00 as compensation for offering their time and travel expenses to participate in each study visit.

## Results

We conducted 6 FGDs with 55 participants consisting of separate sessions for 36 adolescents (12-19 years) and 19 parents/guardians to ensure open dialogue within each group. For the adolescent FGDs, we had 24 females and 12 males whose median age was 17.5 (interquartile range (IQR) 15-18 years). For the parent/guardian FGDs, we had 17 females and 2 males whose median age was 37 years (interquartile range (IQR) 34-46 years).

At baseline (study entry, April to July 2018), we conducted two FGDs with adolescents (N=17) and two FGDs with parents/guardians (N=19); these baseline FGDs are referred to below as A1 through to A4 for the adolescents, and P1 and P2 for the parents/guardians. The two parent/guardian FGDs were conducted in April and June 2018 and the two adolescent FGDs were conducted in July 2018.

At endline (study exit after completing all study visits, August to November 2019), we conducted an additional two FGDs with a different group of adolescents (N=19). Conducting FGDs at these two timepoints helped identify attributes before and after trial participation.

Our analysis using the COM-B framework revealed distinct patterns in how capability, opportunity, and motivation influenced adolescents’ research participation decisions. Within each component, we identified both motivators and barriers that evolved between baseline and endline, highlighting the dynamic nature of participants’ experiences and perceptions. Below, we present these findings organised by COM-B components, tracing changes over time and examining variations between adolescent and parent/guardian perspectives.

### Psychological Capability

The psychological capability component of the COM-B framework encompasses the psychological skills, knowledge, and understanding needed to engage in research participation (24). In our study, this manifested primarily through two key themes: knowledge acquisition as a motivator and lack of awareness and understanding about the research process as a barrier. The evolution of these themes from baseline to endline revealed how participation in research itself could enhance psychological capability, though certain knowledge gaps persisted throughout the study period (13,14).

### Knowledge Acquisition

Knowledge acquisition emerged as a significant motivator for both adolescents and parents/guardians to participate in clinical research. We observed a notable evolution in understanding and appreciation of this factor from baseline to endline.

At baseline, adolescents demonstrated limited understanding of clinical research processes, with some misconceptions.

A1-G Adolescent: “*People think that when we come to Aurum (clinical research unit), we coming to sell blood.*”

A1-A Adolescent: “*If you have the HIV virus, you will be injected for 6 months than maybe after 6 months you will be okay.*”

However, by the endline, adolescent participants expressed appreciation for the opportunity to learn about health, particularly safe sex practices.

A3-A Adolescent: “*I also learned that whenever you want to engage in sexual activity, you should ask questions to your partner. Ask him if he has any of the disease which can lead you to getting the STDs and STIs.*”

A3-C Adolescent: “*I was more eager to know about the health risks of unprotected sex and the workers here are really kind and are really friendly.*”

A4-J Adolescent: “*I learned about many diseases that I didn’t know. I learned to prevent diseases so that they won’t infect me*.”

The desire to understand one’s body and health was particularly strong among adolescents.

A3-J Adolescent: “*I would like to know about dangerous diseases I could get from sexual intercourse.*”

A4-I Adolescent: “*With me, it is because hmm my health, like my safety. I mean to know everything about me, what is going on, how and when.*”

Parents/guardians also recognized the value of knowledge acquisition through research participation. P1-A Parent/Guardian: “*So we make them aware so that they can take part in vaccine trials, make them aware that prevention is better than cure.* “

P1-B Parent/Guardian: “*I don’t think that all adolescents know because if they knew they wouldn’t engage in unprotected sex because most of our children they become pregnant when they are young, they are getting HIV and AIDS when they are still young because they don’t know, which means, they do not know about that risk or not.”*

### Lack of Awareness and Understanding about the Research Process

Lack of awareness and understanding about the research process, including vaccines, clinical trials, objectives, and potential benefits, emerged as another barrier to adolescent participation in clinical research. This theme was present in both the adolescent and parent/guardians FGDs, with some differences between the baseline and endline FGDs.

At baseline, adolescents demonstrated limited understanding about vaccines and clinical trials.

A1-D Adolescent: “*Children at crèche. What injection is that?*”

A1-A Adolescent: “*If am having sex than I would take part in that vaccine trial but if am not having sex than I won’t take part in the vaccine trial.*”

In contrast, the endline adolescent FGDs revealed a greater understanding of vaccines and trials, likely due to their participation in the study. Participants were able to articulate the purpose of the research more clearly.

A4-G Adolescent: “*I just wanted to know what this study is about, to know my status and about other diseases I don’t know about.*”

A4-H Adolescent: “*It was to generally help people who can benefit from the study to get better treatment of any ailments that they might possess or have.*”

However, some confusion still persisted, particularly regarding data security and the research process.

A4-H Adolescent: “*What if in case the information gets stolen?*”

In the parent/guardian FGDs, lack of awareness was also a theme, particularly in terms of understanding the importance of adolescent participation in research. Parents emphasized the need for better communication and education.

P1-A Parent/Guardian: “*I think that…I think it is about…our children who watch TV, all of them right.*”

This quote suggests that parents recognized the influence of media on adolescent decision-making and the need for accurate information about research participation.

### Physical Opportunity

Physical opportunity within the COM-B framework refers to environmental factors that enable or constrain behaviour (24). In our study context, these opportunities manifested through two primary channels: access to healthcare services as a motivator and logistical challenges as a barrier. The interplay between these factors significantly influenced adolescents’ ability to participate in research, with particular emphasis on the practical aspects of engagement (12)(18).

### Access to Healthcare Services

Access to healthcare services and support from research teams emerged as another significant motivator for adolescents and parents/guardians to participate in clinical research. This theme encompasses two distinct but related aspects: the tangible healthcare services provided, and the supportive environment created by the research staff.

Participants often contrasted their experiences in the study with those at local public health clinics, highlighting improved service quality, respect, and confidentiality.

A1-X Adolescent: “*Like at our local clinics (referring to public health clinics) when you get there looking for help, you get that the reception is rude and slow, then they tell you that they going to lunch. So when you come here you know that things like that will not happen. When you get here, they help you.*”

A1-F Adolescent: “*Sometimes when you go to the clinic people at the clinic start talking about you saying that you have AIDS.*”

A1-D Adolescent: “*Privacy. When the nurse draws blood from us and takes the blood to the lab, she does not write our names on the tub.*”

A3-C Adolescent: “*We were taught about different contraceptives to use. The ones that prevent both STI’s and pregnancy and the ones that only protect against pregnancy.*”

Access to sexual health services and contraceptives was seen as a benefit, although some adolescents expressed discomfort in accessing these services from older staff members

A3-X Adolescent: “*To me it was kind of embarrassing…Like when I have to ask for them…Like asking an older person.*”

### Logistical Challenges

Logistical challenges, including practical issues such as transportation and conflicting study and school schedules, were identified as barriers to adolescent participation in clinical research. These challenges were mentioned in both the baseline and endline FGDs, with some differences between the adolescent and parent/guardian perspectives. At baseline, adolescents mentioned transportation and scheduling conflicts as potential barriers.

A1-A Adolescent: “*Something else that would make me not to come is that when the driver comes to fetch me with the Ford Ranger, people will be thinking that I have a “blesser” when they see me get into the Ford Ranger.*”

A1-A Adolescent: “*Another thing is that today we came on a Monday, next time when we have to come here on a Monday, we will not be able to attend our study visits because we will be at school.*”

In the endline adolescent FGDs, logistical challenges were more prominently discussed, particularly the ongoing challenge of balancing study participation with school commitments. Time management and unpredictable study visit durations were also highlighted as concerns

A3-E Adolescent: “*The problem was the time chosen for visits. Like sometimes we came in the morning and like you have a lecture class and you see they clash. You would find yourself having stress about wanting to come…to your visit but you couldn’t.*”

A3-C Adolescent: “*Usually at school we as grade 11 pupils, we sometimes write math tests after school. So I won’t be the only one not writing the test.*”

A3-B Adolescent: “*I wanted to say that during term 1 and term 2 we write tests or subjects after school.*”

A4-I Adolescent: “*Another thing is time maybe let’s say you said you will come at 8 o’clock and you can’t reach and maybe you have planned that around…at least 12 o’clock you will be out, but your things end up not going the way you have planned. We would have stayed here for long hours.*”

In the parent FGDs, logistical challenges were not as prominently discussed, but some parents did mention transportation as a potential barrier suggesting that transportation arrangements and trust in the study staff could influence adolescent participation. To address these logistical challenges, participants in both the adolescent and parent/guardian FGDs offered some suggestions.

A3-E Adolescent: “*Like they call a day before you come like they don’t give you that thing like when you lost that schedule paper.*”

P2-F Parent/Guardian: “*I think that the small children should be picked up at home that way they will not be afraid.*”

These suggestions highlight the importance of reminders, clear communication about study visits, and the need for safe and reliable transportation options.

### Social Opportunity

Social opportunity, as defined in the COM-B framework, encompasses the cultural milieu and social cues that make behaviour possible or prompt it (24). In our study, this manifested through two distinct but interrelated themes: community engagement and support from research teams. These social factors played a crucial role in shaping both the willingness and ability of adolescents to participate in clinical research, particularly through their influence on trust and social acceptance (1)(19).

### Community Engagement

Community engagement emerged as a pivotal theme in the discussions, particularly at baseline, with participants highlighting its potential to foster trust, disseminate information, and ensure the study’s success.

It is important to note that this topic was not explicitly repeated in the end-line FGDs, which is why the results in this section focus almost entirely on baseline data. Despite this, the insights gathered at baseline provide valuable information about the role of community engagement in clinical research participation.

Adolescents expressed a desire for more interactive sessions that would bridge the gap between the research and the community, and even more pertinently, their parents/guardians.

A2-B Adolescent: “*I think in the community, they do not encourage us to do things that are not good, instead they encourage us to do things that are good.*”

Parents/guardians, too, saw the value in leveraging community structures and networks at baseline. They believed that the study’s success hinged on its ability to resonate with the community’s ethos and values.

P1-B Parent/Guardian: “*As a community it needs our participation on teaching our children.*” Parents/guardians also acknowledged their role in determining whether an adolescent went on to participate in clinical research or not as a reflection of community perceptions.

P2-H Parent/Guardian: “*When the child and the parent have an agreement that (then) the child can take part.*”

The role of local leaders, especially elders and community influencers, was frequently highlighted. Parents/guardians believed that their endorsement could significantly influence the community’s perception of the study. The participants’ insights underscored the importance of a community-centric approach. They believed that for the study to be successful, it needed to be deeply rooted in the community, reflecting its values, aspirations, and ethos.

### Support from Research Teams

The supportive environment provided by the research staff was particularly appreciated, especially at the endline. Adolescents highlighted the non-judgmental and supportive attitude of the research staff as a motivator for continued participation. Participants also appreciated the friendly and comfortable environment created by the research staff.

A3-J Adolescent: “*Whenever we feel uncomfortable, they spot you out of the blue and make you feel comfortable by talking to you.*”

A3-A Adolescent: “*If that certain day you came right here in this place and like you were not okay somehow…uhm they kind of like cheer you up and make you smile. That is what I love about the staff here, they are friendly, they are funny and they are so great.*”

Parents/guardians also recognised the value of the supportive environment and the opportunity for open communication about sensitive topics.

P2-B Parent/Guardian: “ *If we do not encourage them as parents, they (adolescents) will not come and they will get wrong information outside. Therefore, we have to encourage them and tell them about Aurum and the importance of coming to Aurum.* “

Parent/Guardian P1-I: “*Sometimes you find yourself being a single parent on girls or boys – so as a parent you will have to be in a position to advice a boy child or a girl child. Tell them the truth. You need to have knowledge so that give you kids right advice.*”

### Reflective Motivation

Reflective motivation in the COM-B framework involves conscious decision-making, plans, and evaluations that drive behaviour (24). In our study, this was primarily expressed through altruistic motivations, where participants demonstrated conscious evaluation of how their participation could benefit others and their community. This reflective process revealed how adolescents and their guardians weighed the broader implications of research participation beyond personal benefit (13,15).

### Altruism

Altruism emerged as a minor, yet notable, motivator for participating in clinical research, particularly through the expressed desire to help and benefit others. This theme demonstrated a recognition that the adolescents’ participation could potentially lead to broader community health benefits.

A1-A: Adolescent: “*Something that will make me become a part of that is that I know that I will also benefit from that and I know that I would have helped many people to get the HIV*

*vaccine. We know that in the communities that we live in, a lot many people are sick, they are not able to get help, and when they go to the clinic, they stand in long lines.*”

In the endline FGDs, altruism was less explicitly mentioned. However, there was one instance where an adolescent participant indirectly referred to altruistic motivations.

A3-A Adolescent: “*The workers working in the Aurum institute do not discriminate the adolescents when they test. Uhm in other words…I mean like visit six and then you are like fine and then you come in for another visit which is in three months’ time and you find that a girl is pregnant or a boy has developed something, uhm they will not judge you basically. They will just…you know support you.*”

This quote suggests participants were willing to continue in the study even if they developed health issues, potentially indicating a form of altruism. However, it is important to note that this interpretation is not explicitly stated by the participant and could also be attributed to other factors such as the supportive environment or personal benefits from continued participation.

### Automatic Motivation

Automatic motivation within the COM-B framework encompasses emotional reactions, desires, and impulses that influence behaviour (24). In our study, this component was primarily evident through emotional responses such as fear and mistrust. These automatic, emotionally-driven responses played a significant role in shaping participation decisions, particularly at baseline, though their influence appeared to evolve over the course of the study (14,17).

### Fear

Fear emerged as a potential barrier to adolescent participation in clinical research, particularly in the baseline FGDs. This theme was less prominent in the parent/guardian discussions and the endline adolescent transcripts.

In the baseline adolescent FGDs, several types of fears were identified. These included fear of judgment or disapproval from friends, as well as boyfriends. There was also a direct mention of fear regarding confidentiality and privacy, especially when discussing sensitive topics like sex.

A1-A Adolescent: “*I think in future when you make appointments for us, we should not come from one area but from different areas because people talk.*”

A2-H Adolescent: “*You will tell your friends that you have joined Aurum then they laugh at you and say that you have HIV.*”

A2-J Adolescent: “*What can prevent us from being part of a study is our boyfriends. They will ask you why are they asking you all those questions, that means that you tell them about our private life and are you sure that they will keep the confidentiality. Things like sex.*”

Additionally, discomfort and lack of trust when discussing sensitive topics was highlighted.

A2-G Adolescent: “*Maybe when you are not comfortable and you feel that you do not trust the people. They will be asking you about sex and you will not feel comfortable with such things.*”

In the endline adolescent FGDs, other persisting fears were associated with clinical processes and potential diagnoses. These ranged from common concerns like fear of needles to more specific worries about discovering health problems.

A3-C Adolescent: “*The most difficult thing was the injections. I’m really afraid of needles.*” A4-D Adolescent: “*It was so painful to take blood*”

A3-A Adolescent: “*The fear of not being sure…You now that you have not being doing it for a while, but the fear of thinking that you will actually find something wrong with your reproductive system.*”

Additionally, some adolescents, particularly female participants, expressed fear of the study interfering with schoolwork and exams.

A3-H Adolescent: “*For me what was difficult was when I was thinking about what time I’m going to do my school work.*”

In the parent/guardian FGDs, fear was mentioned, but it was not as central to the discussion. Parents/guardians were more likely to mention concerns about community perceptions.

P2-G Parent/Guardian “*It could cause fear and also pressure. Sometimes you will discuss these things outside, then those that know something about Aurum, they will be negative, against what you are saying, and then when there are negative against what you guys are telling them, they will start to have fear.*”

### Mistrust

Mistrust of the research system or research organisation emerged as another significant barrier, particularly in the adolescent discussions. This theme was more prominent in the baseline FGDs compared to the endline, suggesting a potential shift in perceptions over time.

At baseline, adolescents expressed concerns about confidentiality, privacy, and the handling of sensitive topics and samples.

A1-G Adolescent: “*People think that when we come to Aurum, we coming to sell blood.*”

A2-G Adolescent: “*Maybe when you are not comfortable and you feel that you do not trust the people. They will be asking you about sex and you will not feel comfortable with such things.*”

In the endline adolescent FGDs, while mistrust was still present, it appeared less central to the discussions. However, some participants continued to express concerns about the potential misuse of their information.

In A4, and endline adolescent FGD, a participant raised a question about the security of their data. While these concerns were not as frequently brought up as in the baseline FGDs., they are worth mentioning. Unlike other concerns which related largely to the current context, the participant voiced misgivings about the security of their data. They were worried that their personal information if shared, could jeopardise the success of future endeavours.

A4-H Adolescent: “*For future reference, I wanted to know if like the information we get, won’t it be used against me? Against me for anything that I would have done and stuff. So, I want to know if any information that will be used here, it would be used against for some person for business ventures or anything in that.*”

This quote highlighted a specific concern about data security and its potential impact on future opportunities, demonstrating a more nuanced understanding of research implications compared to baseline concerns.

Interestingly, some adolescents at the endline expressed increased comfort with the research staff, suggesting a reduction in mistrust over time.

A3-J Adolescent: “*Well I wasn’t sure if I would be able to tell the staff anything that I would be uncomfortable with. For example, I wouldn’t be able to talk to my parents about certain things. So here, I would be able to talk to the staff members.*”

In the parent/guardian FGDs, mistrust was not a prominent theme. However, parents did express concerns about negative community perceptions influencing trust in the research.

## Discussion

The findings of this study provide a nuanced understanding of the motivators and barriers influencing adolescent participation in clinical research within the South African context, specifically in the Northwest Province of South Africa. This research was conducted within the YFS main study, an adolescent observational study that incorporated YFS design and setting. Our results both echo previous research and introduce fresh perspectives that are crucial for designing and implementing future clinical studies in this setting. Through the lens of the COM-B framework, our findings illuminate how capability, opportunity, and motivation interact to shape adolescent participation. The evolution of these components from baseline to endline offers valuable insights for future research design. One of the novel findings of our study is the evolution of participants’ understanding and attitudes from baseline to endline. We purposefully interviewed different participants at our two time-points (i.e., no one was interviewed more than once) however characterising participant perspective at study enrolment and study endline near the end of study follow up offers unique insights into how engagement in research can influence perceptions over time. Notably, endline FGDs revealed more sophisticated understanding of research processes, increased comfort with research procedures, and stronger appreciation for healthcare services compared to baseline discussions, suggesting that exposure to research participation itself can positively influence adolescents’ perceptions and engagement.

The identified motivators and barriers mapped clearly onto the COM-B framework components: psychological capability (knowledge and awareness), physical opportunity (healthcare access and logistics), social opportunity (community engagement), reflective motivation (altruism), and automatic motivation (fear and mistrust). This theoretical grounding helps explain how different factors interact to influence participation decisions. While the COM-B framework has been applied in health behaviour research globally, its specific application to adolescent clinical research participation in African contexts remains limited. Our study represents a novel approach to understanding adolescent research engagement through this theoretical lens, potentially offering a new methodological perspective for future research in similar settings.

Knowledge acquisition emerged as a significant motivator for both adolescents and parents/guardians to participate in clinical research, particularly at the endline. This finding aligns with studies by DiClemente *et al.* (13) and Woolfall *et al.* (18), which identified knowledge acquisition as a key motivator for adolescent participation in clinical trials in both low and middle-income countries (LMICs) and high-income countries (HIC). These insights have critical implications for clinical research recruitment strategies including developing pre-study educational interventions that emphasise knowledge gain, creating age-appropriate educational materials that highlight potential learning opportunities, designing recruitment approaches that explicitly communicate the educational benefits of research participation and incorporating interactive learning components into clinical trial protocols to enhance adolescent engagement.

Access to healthcare services and support from research teams was another prominent motivator. This is consistent with findings reported by Adler (12) and Cunningham-Erves *et al.* (14) in other sub-Saharan African settings. However, our study provides a more detailed exploration of how this motivation evolves over the course of a study, with participants at endline expressing greater appreciation for the non-judgmental and supportive attitude of research staff. This differs from earlier research by highlighting the relational aspects of healthcare service delivery, not just the transactional elements.

Community engagement was highlighted as a pivotal factor in fostering trust, disseminating information, and ensuring the study’s success. While the importance of community engagement has been emphasized by McClure *et al.* (1) and Slack *et al.* (19) in South Africa. Our study provides specific insights into the role of local leaders and community influencers in shaping perceptions of research in the Rustenburg context.

Among the various role-players who could significantly impact a community’s viewpoint on engaging in clinical trials, community leaders emerged as particularly influential in our study. This finding suggests that future research initiatives in similar settings should prioritise engaging with local leadership structures as strategic partners in research design and implementation.

Regarding barriers, our study identified fear, mistrust, lack of awareness, and logistical challenges as significant obstacles to adolescent participation. While these findings are broadly consistent with barriers reported in other sub-Saharan African settings, such as Tanzania (13), Kenya (18), and Uganda (14), our study provides a more nuanced understanding of how these barriers manifest in the specific context of Rustenburg, South Africa.

Notably, the prominence of fear and mistrust as barriers decreased from baseline to endline, suggesting that participation in the study may have contributed to increased trust and awareness among the adolescents. This finding warrants further exploration in future studies and has implications for how researchers approach participant education and engagement over the course of a study.

The implications of our findings for the design and implementation of adolescent-centred clinical research are significant. Researchers should prioritise effective communication strategies, including the use of age-appropriate materials. The evolution of participants’ understanding over time suggests that ongoing education and engagement throughout a study can be beneficial in addressing initial fears and misconceptions.

Understanding these factors through the COM-B framework suggests that interventions should target multiple components simultaneously - enhancing capability through education, creating opportunities through improved accessibility, and addressing both reflective and automatic motivations through community engagement and trust-building. Furthermore, our findings highlight the need for flexible scheduling and transportation solutions to address the logistical challenges faced by adolescent participants, particularly in balancing research participation with school commitments.

Furthermore, our findings highlight the need for flexible scheduling and transportation solutions to address the logistical challenges faced by adolescent participants, particularly in balancing research participation with school commitments. Researchers should consider collaborating with schools and parents to develop strategies that minimize disruption to participants’ education.

While our study has several strengths, including the exploration of both adolescent and parent/guardian perspectives, it is important to acknowledge its limitations. The experiences and beliefs of the adolescent and parent/guardian groups studied may not fully represent the broader spectrum of adolescent experiences across South Africa or the wider region. Additionally, the parent/guardian perspectives were only captured at baseline, limiting our ability to track changes in their views over time.

Future research should consider expanding the demographic and geographical scope to enhance generalizability and capture a more diverse range of perspectives. Additionally, exploring the perspectives of other stakeholders, such as teachers and healthcare providers, could provide a more comprehensive understanding of the factors influencing adolescent participation in clinical research.

### Conclusion

This study offers valuable insights into the complex factors influencing adolescent participation in clinical research in Rustenburg, South Africa. By exploring both motivators and barriers through the COM-B framework, we have uncovered a nuanced picture of how capability (knowledge and understanding), opportunity (healthcare access and logistics), and motivation (altruism, fear, and trust) interact to shape adolescents’ willingness to engage in research.

A key strength of our study lies in its cross-sectional design, which revealed an important finding: participation in the research itself appeared to increase awareness and trust among adolescents over time. This suggests that engagement in research can be a powerful tool for addressing initial barriers and misconceptions.

Our findings have implications to consider for the design and implementation of adolescent-centred clinical research in South Africa and beyond. They underscore the need for age-appropriate, evolving communication strategies throughout the study period, as well as the importance of engaging parents/guardians and building support through trusted community networks. Researchers should also consider developing flexible scheduling and transportation solutions to address logistical challenges, particularly those related to school commitments. Providing comprehensive health education and services as part of the research process emerged as a key motivator and should be prioritized. Additionally, implementing strategies to address fears and misconceptions early in the research process, with ongoing efforts to build and maintain trust, is crucial.

By incorporating these insights, future clinical trials can create a more inclusive and supportive environment that promotes adolescent participation. This is particularly crucial in South Africa, given its large youth population and the pressing need for adolescent-focused health research.

In conclusion, this study contributes significantly to our understanding of adolescent participation in clinical research. By recognizing and addressing the unique motivators and barriers faced by adolescents in specific cultural and socio-economic contexts, we can improve the design and implementation of clinical trials. Ultimately, this will advance the health and well-being of this critical population, paving the way for more effective and inclusive health interventions for young people in South Africa and beyond.

## Supporting Information

- Protocol: An observational prospective study evaluating the feasibility of enrolling adolescents and assessing the uptake of essential health services within an adolescent friendly clinical trial setting
- Parent/legal guardian Focus Group Discussion (FGD) Guide
- Adolescent Focus Group Discussion Guide (FGD) Enrolment Visit
- Adolescent Focus Group Discussion Guide (FGD) Exit Visit

## Data Availability

All relevant data are within the manuscript and its Supporting Information files.

## Acknowledgements

This work was funded by IAVI and made possible by the support of the United States Agency for International Development (USAID). The full list of IAVI donors is available at http://www.iavi.org. The contents of this manuscript are the responsibility of the authors and do not necessarily reflect the views of USAID or the US Government.

